# Comparison of surgical versus medical termination of pregnancy between 13-20 weeks of gestation in Ethiopia: A quasi-experimental study

**DOI:** 10.1101/2020.10.23.20200527

**Authors:** Tesfaye H. Tufa, Sarah Prager, Mekitie Wondafrash, Shikur Mohammed, Nicole Byl, Jason Bell

**Affiliations:** Department of Obstetrics and Gynecology, Saint Paul’s Hospital Millennium Medical College, Addis Ababa Ethiopia; Department of Obstetrics & Gynecology, University of Washington, Seattle, Washington, USA; Department of Public Health, Saint Paul’s Hospital Millennium Medical College, Addis Ababa Ethiopia; University of Michigan Medical School, Ann Arbor, Michigan USA; Department of Obstetrics & Gynecology, University of Michigan, Ann Arbor, Michigan USA

**Keywords:** Dilation and evacuation, Second Trimester, Medical abortion, Ethiopia

## Abstract

**Background:** Dilation and evacuation is a method of second trimester pregnancy termination introduced recently in Ethiopia. However, little is known about the safety and effectiveness of this method in an Ethiopian setting. Therefore, the study is intended to determine the safety and effectiveness of dilation and evacuation for surgical abortion as compared to medical abortion between 13-20 weeks’ gestational age.

**Methods:** This is a quasi-experimental study of women receiving second trimester termination of pregnancy between 13-20 weeks. Patients were allocated to either medical or surgical abortion based on their preference. A structured questionnaire was used to collect demographic information and clinical data upon admission. Procedure related information was collected after the procedure was completed and before the patient was discharged. Additionally, women were contacted 2 weeks after the procedure to evaluate for post-procedural complications. The primary outcome of the study was a composite complication rate. Data were collected using Open Data Kit and then analyzed using Stata version 14.2. Univariate analyses were performed using means (standard deviation), or medians (interquartile range) when the distribution was not normal. Multivariate logistic regression was also performed to control for confounders.

**Results:** Two hundred nineteen women chose medical abortion and 60 chose surgical abortion. The composite complication rate is not significantly different among medical and surgical abortion patients (15% versus 10%; p=0.52). Nine patients (4.1%) in the medical arm required additional intervention to complete the abortion, while none of the surgical abortion patients required additional intervention. Median (IQR) hospital stay was significantly longer in the medical group at 24 (12-24) hours versus 6(4-6) hours in the surgical group p<0.001.

**Conclusion:** From the current study findings, we concluded that there is no difference in safety between surgical and medical methods of abortion. This study demonstrates that surgical abortion can be used as a safe and effective alternative to medical abortion and should be offered equivalently with medical abortion, per the patient’s preference.

## Background

Worldwide, termination of pregnancy is one of the most common procedures for reproductive aged women [1,2]. Abortion is safe when provided by a trained person using recommended techniques [3]. Despite limited evidence in developing countries, the safety of both surgical and medical methods of abortion in the second trimester is well demonstrated worldwide [4]. Both medical and surgical abortion have evolved to improve in effectiveness, acceptability, and rate of complications [5].

In countries with broadly accessible abortion services, most abortions occur in the first trimester [6]. Often, second trimester abortion is the only option available to women for many reasons, including but not limited to poor access to services and fear of disclosure of pregnancy [7–10].

Medical termination of pregnancy involves the use of medications (mifepristone and misoprostol) to cause the uterus to expel the pregnancy while surgical abortion involves preparing the cervix with mechanical or medical methods followed by dilation and evacuation (D&E) using instruments within the uterus (3,10).

Historical and contemporary studies have compared modern medical and surgical methods of termination of pregnancy resulting in a continued debate about the optimal method of second trimester abortion. Many studies show increased safety and effectiveness with surgical abortion, though others show a slight increase in the risk of fever requiring antibiotics with surgical abortion [12–15].

Motivated by the high rate of maternal death associated with unsafe abortion, Ethiopia adopted a legal reform in 2005 which expanded indications for safe abortion service [16]. Some of the indications in the new law include pregnancy as a result of rape or incest, fetal congenital abnormality, presence of physical and mental disabilities for the patient, and others [16]. Currently, the available working document in the country considers dilation and evacuation a second line of management in the second trimester, to be used only when there is a failed medical abortion [17].

Following the recent creation of a family planning fellowship program in a tertiary academic institution, St Paul’s Hospital Millennium Medical College (SPHMMC), started offering both medical and surgical abortion options up to 20 weeks’ gestational age. A review of 46 cases of D&E in the same institution demonstrated clinical safety with an overall complication rate of 2.3% [18]. This method of abortion newly introduced in the Ethiopian setting has a very low uptake, partly due to fear of acute complications and lack of robust evidence demonstrating effectiveness and safety in an Ethiopian context.

Currently, no published data compare medical and surgical abortion in Ethiopia. This study aims to demonstrate the comparative safety and efficacy of medical and surgical abortion between 13-20 gestational weeks.

## Materials and Methods

### Study design and study participants

A non-equivalent comparison group, quasi-experimental design was employed to include patients who were undergoing elective termination of pregnancy from 13-20 gestational weeks between November 1, 2018 and October 31, 2019. The study was conducted at Saint Paul’s Hospital Millennium Medical College (SPHMMC), a tertiary referral and teaching hospital in Addis Ababa, Ethiopia.

### Second trimester safe abortion standard of care in the institution

All patients with a gestational age between 13-20 weeks are fully counseled on the risks and benefits and given the option of medical or surgical abortion. Those who choose medical abortion receive mifepristone 200 mg on day 1 and are appointed to return to SPHMMC 24-48 hours later for admission and misoprostol administration. Those who chose surgical abortion are given mifepristone 200 mg with or without laminaria and appointed to return the next day for surgical abortion. The decision to prepare the cervix with laminaria is made based on the gestational age. Those with advanced gestational age (>16 weeks) receive laminaria on the same day they receive mifepristone.

Gestational age is determined by ultrasound using a composite calculation including the fetal parameters of head circumference, biparietal diameter and femoral length). The ultrasound findings are documented in the clinical chart. During the procedure, both medical and surgical abortion patients receive pain medication.

In the study, all women presenting for abortion care (medical or surgical) were approached by the study team to ascertain willingness to participate in the study. The inclusion criteria for enrollment were, I) gestational age between 13 – 20 weeks confirmed with ultrasound, II) age ≥18 years, III) willingness to provide informed consent for the study, IV) having a working phone and willingness to be contacted for a follow-up questionnaire 2 weeks after the procedure.

The study team provided interested patients with an overview of the study and the consent form, which they signed. Patients were interviewed by trained data collectors in their preferred language. A standardized questionnaire was written in English and translated into a local language (Amharic). Baseline sociodemographic and clinical information were collected upon admission to the hospital. The remaining procedure-related information was collected after they recovered from the abortion procedure.

Enrolled patients were followed throughout their time in the hospital for immediate complications or side effects related to the abortion procedure. Procedure-related information, including the decision for post-procedure observation, need and indication for additional intervention, and family planning related information, was abstracted from the provider’s note in the clinical chart. After discharge, patients were contacted via phone to answer follow-up questions.

The primary outcome of the study was a composite complication rate, defined as the presence of one of the following complications: bleeding requiring observation, genital tract lacerations requiring repair, need for additional intervention to complete the abortion procedure, and one or more symptoms of pelvic infection at the time of follow up. We also assessed for the presence of other serious maternal complications including continued bleeding for more than two weeks, additional major surgery (laparotomy), and death. Also assessed were duration of hospital stay, degree of pain related to abortion procedure, acceptability of the procedure (as measured by patient satisfaction) and recommendation of the procedure to others. We based our sample size calculation on the composite complication rate of a previous cross-sectional study conducted in the same institution [18]. We assumed a clinically meaningful difference in composite complication rate of 15% between the groups with 80% power and a two-sided alpha of 0.05. Considering the low uptake of surgical abortion in the institution, we planned for a 3:1 ratio of medical versus surgical abortions. Given these assumptions, we needed at least 54 surgical and 162 medical abortion cases. Based on prior numbers of abortions performed in the hospital, we anticipated a one-year data collection would be sufficient to meet our sample size requirements.

The data were collected with open data kit then imported into Stata version 14.2 for data processing and analysis. Univariate analyses were performed using proportions and means (standard deviation), or medians (interquartile range) when the distribution was not normal. The different outcomes were compared between medical and surgical abortion procedure groups using Fisher’s exact test, independent t-test, or the non-parametric test of difference of means (Mann Whitney U test). A multiple logistic regression model was fitted to determine independent predictors of post-procedure complications while adjusting for measured confounding variables.

## Results

During the study period, a total of 279 women were included in the study, with 219 in the medical group and 60 in the surgical group (Figure.1).

**Figure 1.**
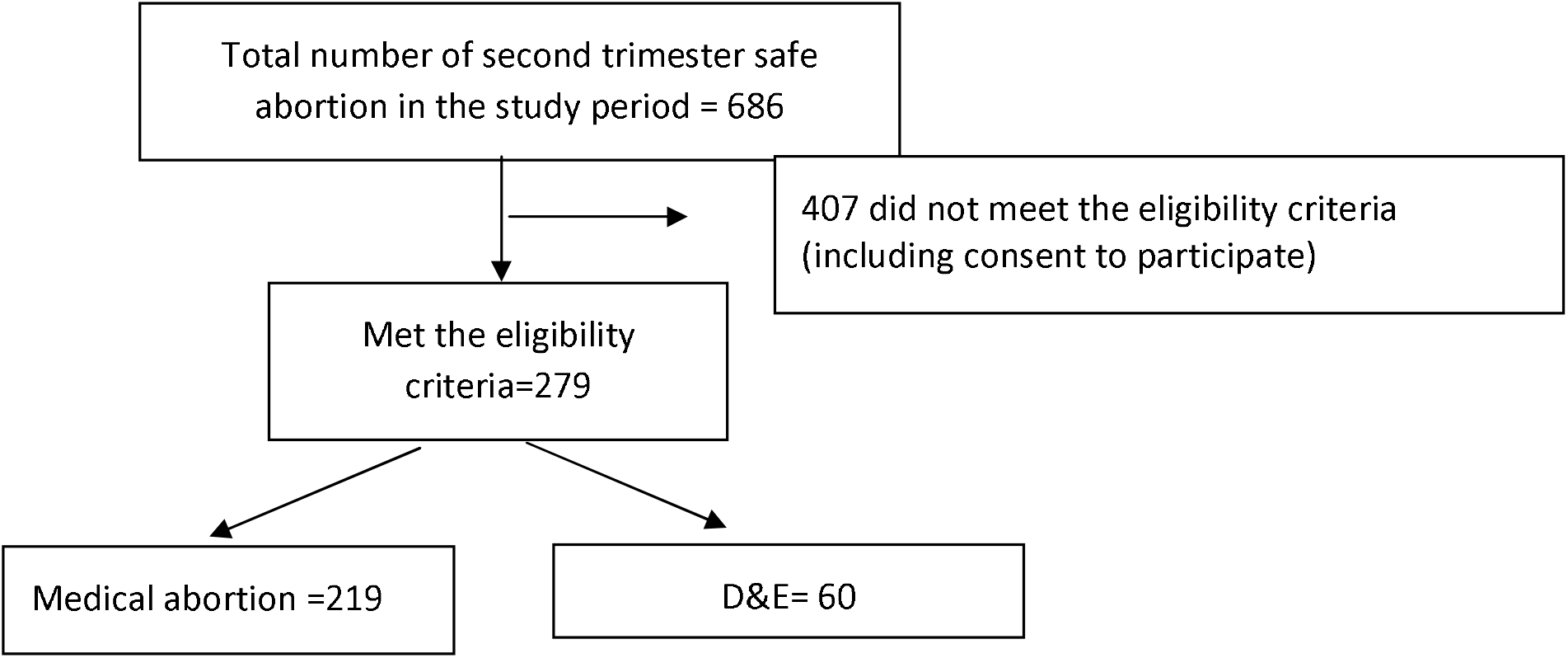
Flow chart of patient enrollment.

The baseline demographic characteristics for the two groups are similar except for age, place of residence, and prior delivery (Table 1). Significantly more patients in the surgical arm are older, live outside of Addis Ababa, and have had a prior vaginal delivery (p <0.05). Other clinical characteristics, including gestational age, were similar between the groups.

**Table 1.** Baseline characteristics of study participants.

| **Characteristic** | **Level** | **Medical**<br>**(n=219)** | **Surgical**<br>**(n=60)** | **p-value** |
| --- | --- | --- | --- | --- |
| Age | 18-25 | 176 (80.4%) | 38 (63.3%) | 0.008 |
|  | 26-35 | 41 (18.7%) | 19 (31.7%) |  |
|  | >35 | 2 (0.9%) | 3 (5.0%) |  |
| Religion | Muslim | 34 (15.5%) | 8 (13.3%) | 0.92 |
|  | Orthodox | 146 (66.7%) | 41 (68.3%) |  |
|  | Protestant | 39 (17.8%) | 11 (18.3%) |  |
| Income, median (IQR) |  | 1000 (700, 2500) | 1500 (1000, 2750) | 0.14 |
| Current residence | Addis Ababa | 176 (80.4%) | 38 (63.3%) | 0.006 |
|  | Outside Addis Ababa | 43 (19.6%) | 22 (36.7%) |  |
| Accompanying partner | No | 180 (82.2%) | 43 (71.7%) | 0.071 |
|  | Yes | 39 (17.8%) | 17 (28.3%) |  |
| Level of education | No formal education | 18 (8.2%) | 6 (10.0%) | 0.42 |
|  | Grade 1-8 complete | 91 (41.6%) | 19 (31.7%) |  |
|  | Grade 8-12 complete | 78 (35.6%) | 22 (36.7%) |  |
|  | College graduate | 32 (14.6%) | 13 (21.7%) |  |
| Gestational age (weeks) | 13-15.6 | 68 (31.1%) | 26 (43.3%) | 0.20 |
|  | 16-17.6 | 47 (21.5%) | 11 (18.3%) |  |
|  | 18-20.6 | 104 (47.5%) | 23 (38.3%) |  |
| Previous abortion | No | 205 (93.6%) | 52 (86.7%) | 0.077 |
|  | Yes | 14 (6.4%) | 8 (13.3%) |  |
| Prior delivery | No | 160 (73%) | 33 (55%) | 0.008 |
|  | Yes | 59 (27%) | 27 (45%) |  |

All patients in the medical arm received mifepristone 24-48 hours prior to misoprostol. Two hundred fifteen patients (98.2%) received misoprostol 400 micrograms every 3 hours, while four patients (1.8%) received a loading dose of 800 microgram misoprostol, then 400 micrograms every three hours. Sublingual was the most common route of administration for misoprostol (88.6%).

Ninety-eight percent (59/60) of patients in the surgical arm received mifepristone on day one. In addition to the mifepristone, surgical patients either received laminaria only (13.3%), misoprostol only (56.7%), or both laminaria and misoprostol (30.0%) for cervical priming prior to dilation and evacuation.

One hundred forty-four women in the medical arm (65.8%) received analgesia. Diclofenac and Ibuprofen were the most common analgesics used for medical abortions, accounting for 50% and 36.8% respectively. Other analgesics used were tramadol and paracetamol. All patients in the surgical arm received a paracervical block, and 53 (91.4%) additionally received diclofenac. Other pain medications used for surgical abortion were Diazepam and Pethidine (Table 2).

**Table 2.**
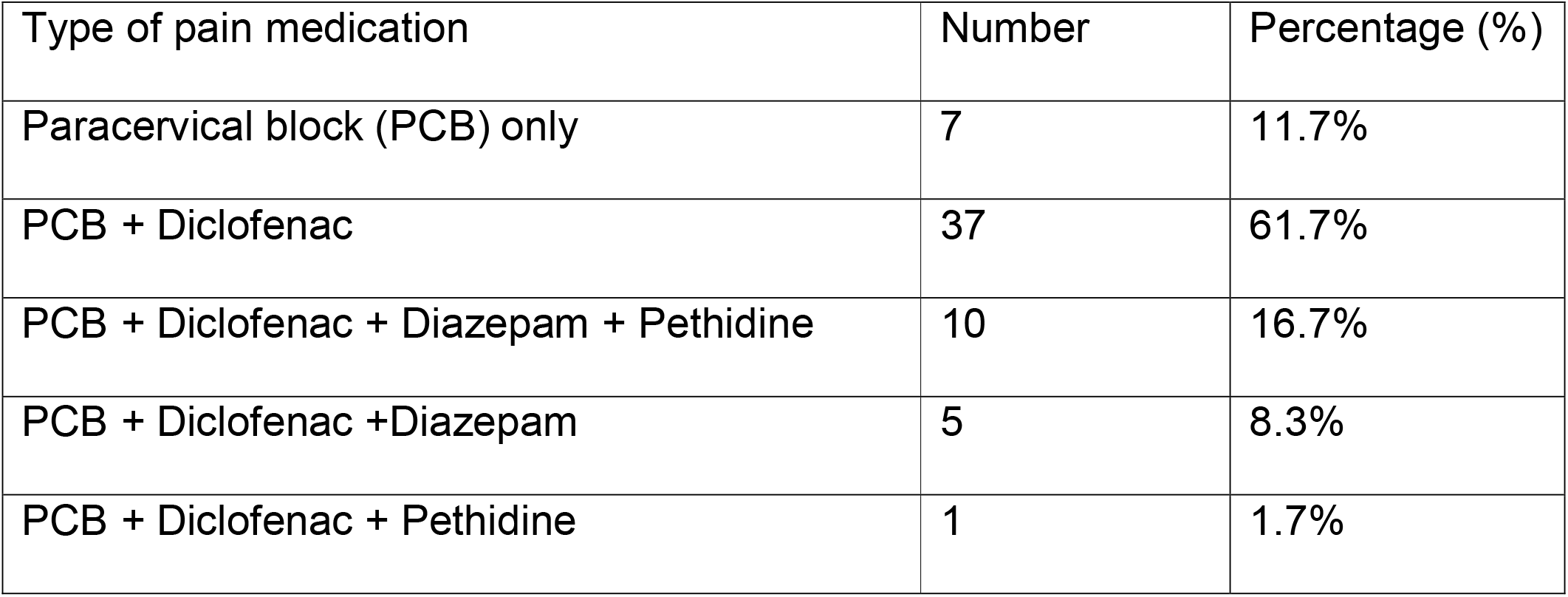
Pain medication used during surgical abortions.

The composite complication rate among medical abortion patients is not significantly different than that of surgical abortion patients (15% versus 10%; p=0.52). Nine patients (4.1%) in the medical arm required additional intervention (manual vacuum aspiration) to complete the abortion, while all patients in the surgical arm completed the procedure without a need for additional intervention. More patients in the surgical arm were observed for bleeding (8.3% versus 1%; p< 0.002). Despite this, no patients in either arm required transfusion or any other medical or surgical intervention to control bleeding. There were no genital tract lacerations requiring repair in either arm and no maternal deaths reported during the study period.

We considered possible confounders and performed multivariate logistic regression to compare the composite complication rate between the medical and surgical arms and found no significant difference between the groups after controlling for measured confounders (Table 4).

**Table 3.** Procedure details and complications of study participants.

| **Factor** | **Medical**<br>**(n=219)** | **Surgical**<br>**(n=60)** | **p-value** |
| --- | --- | --- | --- |
| Composite complication | 33 (15%) | 6 (10%) | 0.52 |
| Observed for bleeding | 1 (0.5%) | 5 (8.3%) | 0.002 |
| Symptoms of pelvic infection | 21 (9.6%) | 1 (1.7%) | 0.055 |
| Continued bleeding for two weeks | 84 (38.3%) | 21 (35%) | 0.65 |
| Duration of hospital stay (hours), median (IQR) | 24 (12,24) | 6 (4,6) | <0.001 |
| Pain scale-medical-surgical |  |  |  |
| No pain | 0 | 5 (8.3%) | <0.001 |
| Mild pain | 48 (21.9%) | 42 (70.0%) |  |
| Moderate pain | 117 (53.4%) | 11 (18.3%) |  |
| Severe pain | 54 (24.7%) | 2 (3.3%) |  |
| Would recommend to others | 184 (84.0%) | 59 (98.3%) | 0.003 |

**Table 4.** Multiple Logistic regression analysis of composite complication.

| **Variables** | **Composite Complication\*** | **Adjusted OR** | **95% CI** |
| --- | --- | --- | --- |
|  | **N (%)** |  |  |
| **Method of abortion** |  |  |  |
| Medical | 30 (83.3%) | 1.00 |  |
| Surgical | 6 (16.7%) | 0.46 | 0.16-1.3 |
| **Age** |  |  |  |
| 18-25 | 27 (75.0%) | 1.00 |  |
| 26-35 | 8 (22.2%) | 1.44 | 0.49-4.2 |
| >=36 | 1 (2.8%) | 2.64 | 0.22-32.2 |
| **Gestational age** |  |  |  |
| 13-15.6 | 20 (55.6%) | 1.00 |  |
| 16-17.6 | 4 (11.1%) | 0.22 | 0.07-0.72 |
| 18-20.6 | 12 (33.3%) | 0.37 | 0.17-0.82 |
| **Previous abortion** |  |  |  |
| No | 32 (88.9%) | 1.00 |  |
| Yes | 4 (11.1%) | 2.39 | 0.6-9.1 |
| **Parity** |  |  |  |
| 0 | 26 (72.2%) | 1.00 |  |
| 1 | 3 (8.3%) | 0.26 | 0.06-1.1 |
| >=2 | 7 (19.4%) | 1.01 | 0.32-3.2 |

\*composite complication (observation for bleeding, one or more symptoms of infection, additional procedures required to complete the abortion). Those having one or more symptoms are considered as having complication (coded as; Yes=1, No=0).

Medical abortion was associated with a significantly longer median hospital stay (24 hrs., IQR 12-24 hrs.) as compared to surgical abortion (6 hrs., IQR 4-6 hrs.; p< .001). The surgical patients were more likely than the medical patients to recommend the same procedure to others (98.3% vs 84.0%; p=0.003).

All patients in both the medical and surgical arms were counseled on family planning and 92% (257/279) received some form of family planning. The majority of the patients chose and received long acting reversible methods, with Nexplanon, IUD and Jadelle contributing nearly 86% of the total contraception methods adopted (Figure 2).

**Figure. 2.**
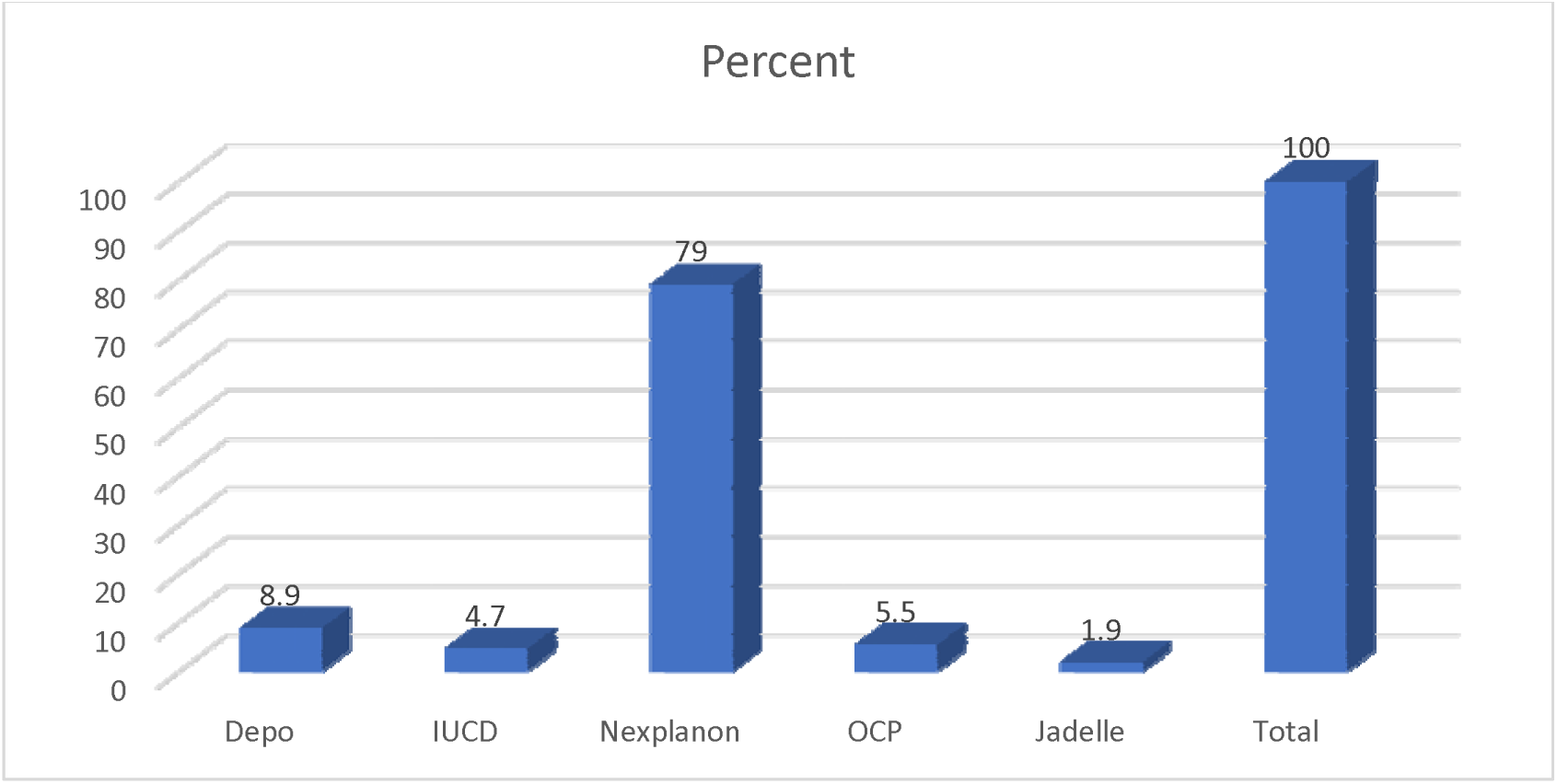
Post abortion family planning.

## Discussion

The low complication rate in our study demonstrates that both medical and surgical abortion between 13-20 weeks can be safely practiced in an Ethiopian setting. Similar to other studies, the need for additional interventions to complete the abortion, occurred more frequently in the medical group [8,14,15,19–21]. At the time of the 2-week follow up enquiry, both ongoing bleeding and symptoms of pelvic infection were similar between the two groups.

All surgical abortion patients received prophylactic antibiotics and some form of pain medication during the abortion procedure. Conversely, only 65.8% of medical abortion patients received pain medication, which may explain the higher reports of moderate to severe pain in the medical as compared to the surgical arm. This reveals the need for standardization of pain management, especially for medical abortion. Additionally, further investigation is needed to identify the reasons for lower pain medication administration among women undergoing a medical abortion.

The median duration of stay is significantly shorter in the surgical abortion group. This finding is consistent with other studies [11,14]. Interestingly, patients who came from outside Addis Ababa were more likely to choose the surgical procedure, possibly to avoid the prolonged hospital stay associated with medical abortion. Similar to other studies, the mean satisfaction rate (scored from 0-8) between the groups is equivalent (7.56 vs 7.90; p=0.995) (3, 19–21). Importantly, this study found that 42.3% of all patients were referred from nearby health institutions. As our hospital is a center of excellence for abortion care, it is good that nearby institutions know where/how to refer patients for abortion. Alternatively, this also indicates a need for capacity building at nearby health centers to reduce/avoid unnecessary delays in receiving care. Integrating D&E training into medical schools and residency programs is one way to build capacity and potentially address these delays. Finally, the high rate of family planning counseling and provision demonstrates the successful integration of family planning into abortion care at the institution.

### Strengths and limitations

This study is the first in Ethiopia to compare and report the safety and efficacy of surgical versus medical abortion. The study enrolled adequate numbers of patients in the medical and surgical arms during the study period. Saint Paul’s Hospital Millennium Medical College is the only public institution so far in the country providing D&E for abortion patients, and only recently started this practice, together with the initiation of a family planning fellowship for graduates of an OBGYN residency program. This study demonstrates that D&E can be safely and effectively initiated and practiced in other similar Ethiopian setttings.

The study has several limitations. Study subjects were not randomized to abortion methods, which introduces selection bias, as revealed by the proportion of patients from outside Addis Ababa who chose surgical abortion. Our study limited the comparison to cases between the gestational age of 13-20 weeks and cannot be generalized to abortion before 13 or beyond 20 weeks’ gestation. The study is performed in an established academic institution where there is a family planning fellowship program and should not be generalized to other hospitals that lack sufficient training and oversight in surgical abortion. Significantly fewer patients chose surgical abortion during the study period. Further investigation into reasons for this is needed to identify knowledge gaps and make recommendations.

## Conclusion

Our study demonstrates that surgical abortion is a safe and effective alternative to medical abortion in our institution. We hope these results will inform the current national technical guidelines, which currently consider second trimester surgical abortion a backup method only. Instead, surgical abortion should be a first line option for patients, along with medical abortion.

Attempts to expand the family planning fellowship program and train more providers in Ethiopia to perform surgical abortion should be part of any strategy to improve care.

## Data Availability

All the relevant data are included in the manuscript

## Acknowledgment

Our deepest appreciation goes to SPHMMC nurses and midwives working in the abortion procedure room who devoted their time and energy to a continued quality of care for abortion patients. We also want to thank SPHMMC’s Center of Excellence in Reproductive Health for supporting the research. We deeply appreciate Dr. Soe Soe Thwin for her revisions and technical input.

## Funding statement

The research received grant from Saint Paul’s Hospital Millennium Medical College, Addis Ababa, Ethiopia.

## Competing Interest

The authors declare no competing interests.

## Consent for publication

Not applicable

## Availability of data and materials

The primary dataset used in the study is not publicly available. In case it is needed, the data can be obtained from the corresponding author upon reasonable request.

## Contribution of the authors

TT: Conceived and conducted the study, managed the data, performed the statistical analysis and write up the manuscript, JB: Overall supervision for the entire process of study planning, implementing, manuscript writing, MW: Statistical analysis, data interpretation, Critical revision of the manuscript. SP: Overall supervision for the entire process of study planning, implementing, manuscript writing, SM: Overall supervision for the entire process of study planning, implementing, manuscript writing, NB: Critical revision and Manuscript editing

## Ethics approval and consent to participate

Ethical approval was received from the ethical review board at SPHMMC and permission to perform the research was obtained from the head of the department of obstetrics and gynecology. The study was conducted after receiving an official letter of clearance from SPHMMC ethical review committee.

Informed consent was obtained from all the surveyed women prior to the interview.

## Notes

### Competing Interest Statement

The authors have declared no competing interest.

### Clinical Trial

NCT04602052

### Author Declarations

Ethical approval was received from the ethical review board at SPHMMC and permission to perform the research was obtained from the head of the department of obstetrics and gynecology. The study was conducted after receiving an official letter of clearance from SPHMMC ethical review committee. Informed consent was obtained from all the surveyed women prior to the interview.

